# Spatio-temporal analysis between the incidence of COVID-19 and human development in Mato Grosso do Sul, Brazil

**DOI:** 10.1101/2021.01.19.21250106

**Authors:** Amaury de Souza, Marcel Carvalho Abreu, José Francisco de Oliveira-Júnior

## Abstract

**Objetive:** To analyze the spatial distribution of the Covid-19 incidence and its correlation with the municipal human development index (IDHM) in the state of Mato Grosso do Sul (MS), Brazil.

**Methods:** This is an ecological, exploratory and analytical study whose units of analysis were the 79 municipalities that make up the state of MS. Covid-19 incidence coefficients, death numbers, lethality rate, mortality rate and Human Development Index for municipalities (IDHM) in the period from March 2020 to December 31, 2020 were used. spatial correlations between the variables mentioned above.

**Results:** The incidence of Covid-19 has spatial dependence with moderate positive correlation and formation of clusters located in the Metropolitan Region of Campo Grande (RMCG) and municipalities in the region.

**Conclusion:** The uneven mapping of Covid-19 and its relationship with IDHM in the Ministry of Health can contribute to actions to address the regional pandemic.

## Introduction

On December 31, 2019, China reported to the World Health Organization (WHO) that a group of individuals was diagnosed with a type of pneumonia, with no defined etiology, detected in the city of Wuhan, Hubei province in China (1-4). After 12 days, the Chinese shared the sequencing of the new virus, the Severe Acute Respiratory Syndrome Coronavirus 2 (SARS-CoV-2) (2,4). In the same month, WHO convened two meetings of the Emergency Committee, the last of which noted the growth in the number of cases and countries affected, and thus declared the situation as a Public Health Emergency of International Importance (ESPII) (3, 4).

A worldwide systematic review evaluated the epidemiological profile of Covid-19 and found that the mortality rate for this cause reaches 0.3% of the population, being higher in elderly men (5).

In Brazil, the first case of the Coronavirus disease (COVID-19) was confirmed on February 26, 2020, in the city of São Paulo (5) and in Campo Grande on March 14, 2020.

According to studies, the average time for the disease to double varies between 5.2 to 7.4 days (6-8). Thus, the speed of progression of the epidemic is high. Given this potential for disease transmission, along with the absence of immediate measures capable of containing the infection, such as drugs or vaccines, interventions in the social structure are essential and urgent measures. In this sense, social isolation proved to be the most appropriate option for these conditions, mainly due to the support of world experiences. The measures of social distance aim to avoid the overcrowding of the Unified Health System (SUS), it is one of the largest and most complex public health systems in the world, which would occur through simultaneous and massive infection, in order to guarantee adequate access to the population. ventilatory support and care in Intensive Care Units (UTI) (6).

However, it is known that the economic impact caused by the pandemic and the paralysis of several commercial activities is great, especially in countries considered to be underdeveloped or developing, with important consequences on the population’s quality of life (9-11). In Brazil, the need to adopt periods of greater flexibility in social isolation measures is frequently debated, aiming to guarantee a source of income and economic movement in the country (10). For this to be possible, the effective action of Epidemiological Surveillance is essential. SUS recommends that this surveillance be carried out, that is, actions oriented towards the effective collection of data, in order to define the dynamics of a disease or health problem. With the variables collected, epidemiological information is produced and disseminated, which serve to guide the management of the various services and levels of care (12,13). In the pandemic scenario, this epidemiological monitoring plays a fundamental role in coping with the disease, since the analysis of the data allows the verification of the patterns referring to COVID-19 in order to predict its behavior, as well as its evolution. In this way, important information is generated for the rational direction of public policies, in the sense of establishing a strategic planning for health surveillance. (6,12,13)

The distribution and diffusion of infectious diseases is an explicitly spatial process (Cordes and Castro, 2020) (14) and cluster analysis (CA) is an important surveillance and planning tool (Andersen et al., 2021) (15) to combat the spread of COVID-19, as well as in strategies to reduce the mortality / lethality of the virus. The CA technique has already been useful in determining spatial patterns of pandemian China (Li et al., 2020) (16), United States (Cordes and Castro, 2020; Andersen et al., 2021; Holh et al., 2020) (14-15-17) and India (Das et al., 2021) (18), in highly densely populated countries in the world. In Brazil, studies with a statistical approach to the spatial patterns of the COVID-19 pandemic are scarce. And the importance of this type of approach is extremely important at a time when the number of cases and deaths is increasing significantly

The state of Mato Grosso do Sul is located in the central-western region of Brazil, and has an area of approximately 358159 km2 (4.19% of the national territory) distributed in 79 municipalities. According to Silva et al. (2020) (19), 73.41% of the state’s municipalities are classified as very vulnerable to COVID-19, especially the municipalities with the largest population.

Thus, the present study aims to: i) analyze the epidemiological profile of COVID-19 in MS and ii) evaluate the spatial distribution of the incidence of Covid-19 and its correlation with the municipal human development index (IDHM).

## Methodology

It is an ecological, exploratory and analytical study whose units of analysis were the 79 municipalities that make up the state of MS. The Covid-19 incidence coefficients in the municipalities in 2020 were used. The social information of the municipalities was collected in the Human Development Atlas in Brazil (20), systematized by the 2010 United Nations Development Program (PNUD). official estimate of the population of each municipality in the state of MS, as well as of that federative unit, was obtained for the year 2020 by consulting the system of the Informatics Department of the Unified Health System (DATASUS) / Health Information (TABNET) (21).

The incidences of Covid-19 were obtained based on the confirmed cases made available by the Health Department of the State of Mato Grosso do Sul through the SUS electronic platform. To calculate the incidence coefficient of each municipality with cases, the number of confirmed cases was divided by the resident population and multiplied by the population base of 100 thousand inhabitants.

As an independent variable, social development was used, with the IDHM variable estimated for each municipality in 2010, as available from PNUD. The IDHM can be classified as very low (0 to 0.499), low (0.500 to 0.599), medium (0.600 to 0.699), high (0.700 to 0.799) and very high (0.800 to 1). The IDHM summarizes in an average of three sub-indices, calculated on the basis of a few indicators easily collected in different nations, three basic and universal dimensions of life, which are the conditions for the choices and opportunities of individuals to be expanded: access to knowledge (education), the right to a long and healthy life (longevity) and the right to a decent standard of living (income). Due to its simplicity, the IDHM does not deepen any of these dimensions particularly, but it allows to compare the general level reached by the units of analysis in meeting these basic needs so that individuals can develop their capacities and their choices (22).

Descriptive statistics was performed with data on the cases and population of each municipality, and the incidence coefficient was then calculated. The IDHM variable was presented by the respective values of the arithmetic mean for the three sub-indices.

All data used were secondary, without personal identification and in the public domain, which, according to Resolution No. 510/2016, of the National Health Council, dispenses with the need for prior approval by the Ethics in Research with Human Beings Committee (23).

The analyzed data refer to the cases reported in the period from March 14, 2020 to December 31, 2020. In this study, the cases that met the diagnostic criteria for COVID-19 were included, according to the guidelines of the Ministry of Health. The variables analyzed were: number of confirmed cases and deaths; number of suspected cases and deaths; number of tests; total number of hospitalized patients (in ICU and infirmary); biological sex (male and female) and age group (<1, 1-9, 10-19, 20-29, 30-39, 40-49, 40-59, 60-69, 70-79 and 80 + years).

The analysis of the number of accumulated cases, the number of new cases, the number of accumulated deaths and the number of new deaths was performed by graphing these variables as a function of time (in days) and first and second degree linear regression analysis (Eqs 2 and 3) and non-linear of the exponential type with 1 and 2 (Eqs. 4 and 5 parameters). The significance of the regression parameters was tested by the Student’s t test and the adjustments were evaluated by the coefficient of determination (r^2^), Willmott’s agreement index (d) and the root of the mean square error (RMSE).

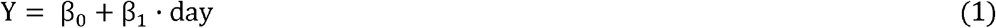

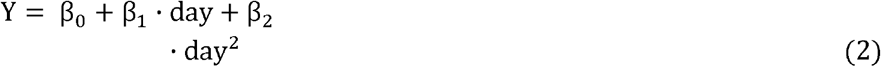

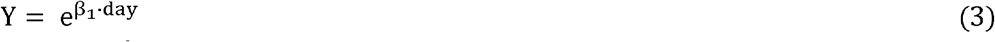

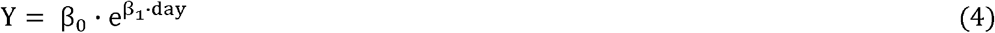

For new cases and new deaths, the 7-day moving average for the period under analysis was calculated and the modified Mann-Kendall (MK) test was applied to verify whether the fluctuations around the average show trends. The MK test is a non-parametric trend verification test that consists of testing the null hypothesis of stationarity in the time series (Gois et al., 2020) (24). For a time series, the Mann-Kendal (S) statistic is defined by:

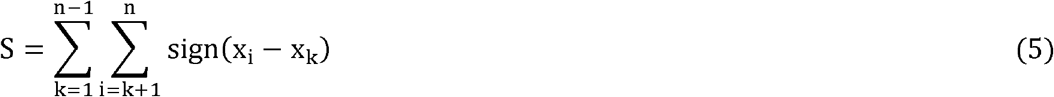

Where *x*_*i*_ is the i-th observation, *x*_*k*_ is the k-ésima observation, immediately after the i-th observation and n is the number of elements in the time series. The arrangement of the signs is given by:

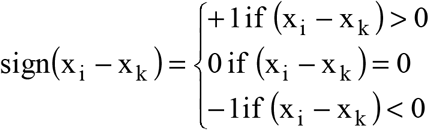

The variance is computed as:

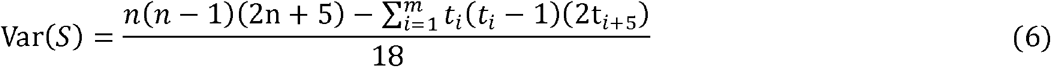

Where m is m is the number of groups tied, *t*_*i*_ is the number of loops of length equal to i.

The MK test requires that the sample data be serially independent (absence of auto correlation). Sample data correlated in series affects the test’s ability to correctly assess the significance of the trend. To eliminate the effect of the serial correlation in the MK test, an autocorrelation analysis was performed, which was performed through the correlogram analysis, which consists of a graph of the autocorrelation function for sequential values of delay k = 0, 1, 2,. ., n. In case of verified autocorrelation, the Mann-Kendall test was applied with the modification proposed by Yue and Yang (2004) (25). The modification by Yue and Wang (2004) (25) consists of a correction of variance to address the issue of serial correlation in the trend analysis, maintaining the significance of the test (Blain, 2013; Sa’adi et al., 2019) (26, 27).

Pettitt’s statistics account for the number of exceedances in which the value of the first sample exceeds that of the second sample. The null hypothesis of the test is that there is no breaking point in the data series (Gois et al., 2020) (24).

The CA technique took into account the variables: average IDH, covid incidence rate, deaths, lethality (%) and mortality, in the cities of the state of MS. The measure of dissimilarity considered was the Euclidean distance (eD), according to Eq. 7:

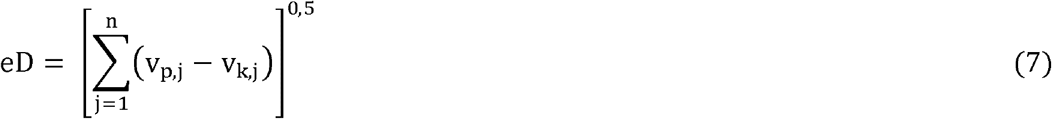

where eD is the Euclidean distance (dimensionless), *v*_*p,j*_ and *v*_*k,j*_ are the quantitative characteristics of the cooperatives of the variable *p* and *k*.

The number of groups was determined by the approach of the average silhouette measures the quality of a cluster by determining how well each object is in its cluster. A high average silhouette width indicates good grouping. The average silhouette method calculates the average silhouette of observations for different numbers of groups (k). The optimal number of k clusters is that which maximizes the average silhouette over a range of possible values for k. The criterion identified 4 groups as the optimal number of clusters for MS.

The hierarchical method of Ward (Ward, 1963) (28) was used for agglomeration, due to the success of this method in studies that aim to obtain similar groups, mainly in biometeorological and collective health studies (Oliveira Júnior et al., 2019 ; Holh et al., 2020; Shariati et al., 2020)(29, 17, 30).

Principal component analysis (ACP) was applied to the same variables in the grouping to verify what most influences the variance of data related to COVID-19 in the state MS. The variables were considered the variables average IDH, incidence rate covid, deaths, lethality (%) and mortality. ACP was performed to explain the structure of data variance through linear combinations of the original variables. The main components were extracted using the correlation matrix between the studied variables.

Before proceeding with PCA, the Bartlett test (B) was performed to test the null hypothesis (H_0_) that the variables are not correlated (there is non-collinearity between the variables), which would make PCA impossible because it depends on the construction linear combination of variables. The Kaiser-Meyer-Olkin index (KMO) was also used to check the suitability of ACP, with a KMO value> 0.5 (Fraga et al., 2020) (31) being desirable.

## Results

Comparatively, in the period from March 14, 2020 to December 31, 2020, in the world we had 83,192,664 cases with 1,813,087 deaths, in Brazil there were 7,675,973 cases with 194,949 deaths and in the state of MS with 133,761 cases with 2,329 deaths with an average lethality rate of 1.7% with 47.4% of cases positive for males and 52.6% of cases for females. In this period, a total of 130,510 tests were performed, with 54.1% RT-PCR (Reverse-Transcriptase Polymerase Chain Reaction) test with 29% positivity, 42.8% rapid tests with 5.3% positivity, 1.9% serology and 1.1% was not informed of these 11,224 were positive (25.11%) and 33,475 were negative (74.89%). The state of MS ranks 4th in notification rates by Brazilian states. However, the number of confirmed cases of COVID-19 in Brazil has been largely underreported, where the actual number of cases may be about 11 times greater than those currently reported (Prado et al. (2020) (32).

The situation of COVID-19 cases in MS were 133,761 confirmed cases, in household isolation 14,382, 116,280 recovered and 2,329 deaths; 670 cases were hospitalized, 360 in clinical beds, 239 in public beds and 121 in private beds; 310 were in the ICU, 220 in public and 90 in private.

What calls attention at COVID-19 are the high rates of allocation of patients in the UTI, corresponding to 45% of daily admissions. However, this high number also reflects the concentration of testing in more severe individuals when compared to WHO data, which indicate that 15% of patients with COVID-19 require hospitalization with oxygen therapy and 5% need to be admitted to the UTI (Noronha et al, 2020) (33).

In this sense, the occupation of UTI beds is linked to admission due to critical instability, with the need for life support intervention, being the priority for patients with a high probability of recovery (Moreira, 2020) (34).

It is noteworthy that the practice of adequate Health Surveillance accompanied by effective epidemiological investigation during epidemics is a mandatory activity of any local health surveillance system and must occur in an integrated manner and concurrently with prevention and control actions, with the dissemination of data with as soon as possible to related services, to higher levels and to society and its representatives (Henrique and Vasconcelos, 2020) (35). In this sense, according to the National Contingency Plan for Human Infection by the new Coronavirus of the Ministry of Health, it is up to the Health Surveillance services to establish risk communication strategies, with the collection and dissemination of information that help in combating and controlling disease (Ministry of Health, 2020) (36). In view of this, the need is reinforced, in times of pandemic, to improve the notification systems and the transfer of information between different levels, as well as to invest in strategies for testing the population associated with monitored social distance policies, with a survey of local social isolation rate, as, as observed in the present study, municipal decrees of commercial closure, without adequate surveillance, may not be sufficient in the control of cases, deaths and hospitalizations.

The age profile of confirmed cases by age group were: (<1 (0.6%), 1-9 (3.0%), 10-19 (6.6%), 20-29 (20.2%), 30-39 (24.3%), 40-49 (19.1%), 40-59 (13.5%), 60-69 (7.6%), 70-79 (3.4%) e >80 (1.7%)).

For the cumulative number of COVID-19 cases and deaths, the periods with the highest rates of increase in 2019 occur between the beginning of July and October 20, and from November (Figure 1). It is worth mentioning that in this period, municipal elections took place throughout Brazil, which should not have been held. A slight decrease in the rate of increase in cases of death was observed between the end of September and mid-November. The new cases and new deaths, on the other hand, had a similar behavior over time, in which significant peaks were observed between the month of July and October, and between November and December, in the case of the month of December, fraternizations and agglomerations for purchases in commerce are due, common at this time of year.

**Figure 1.**
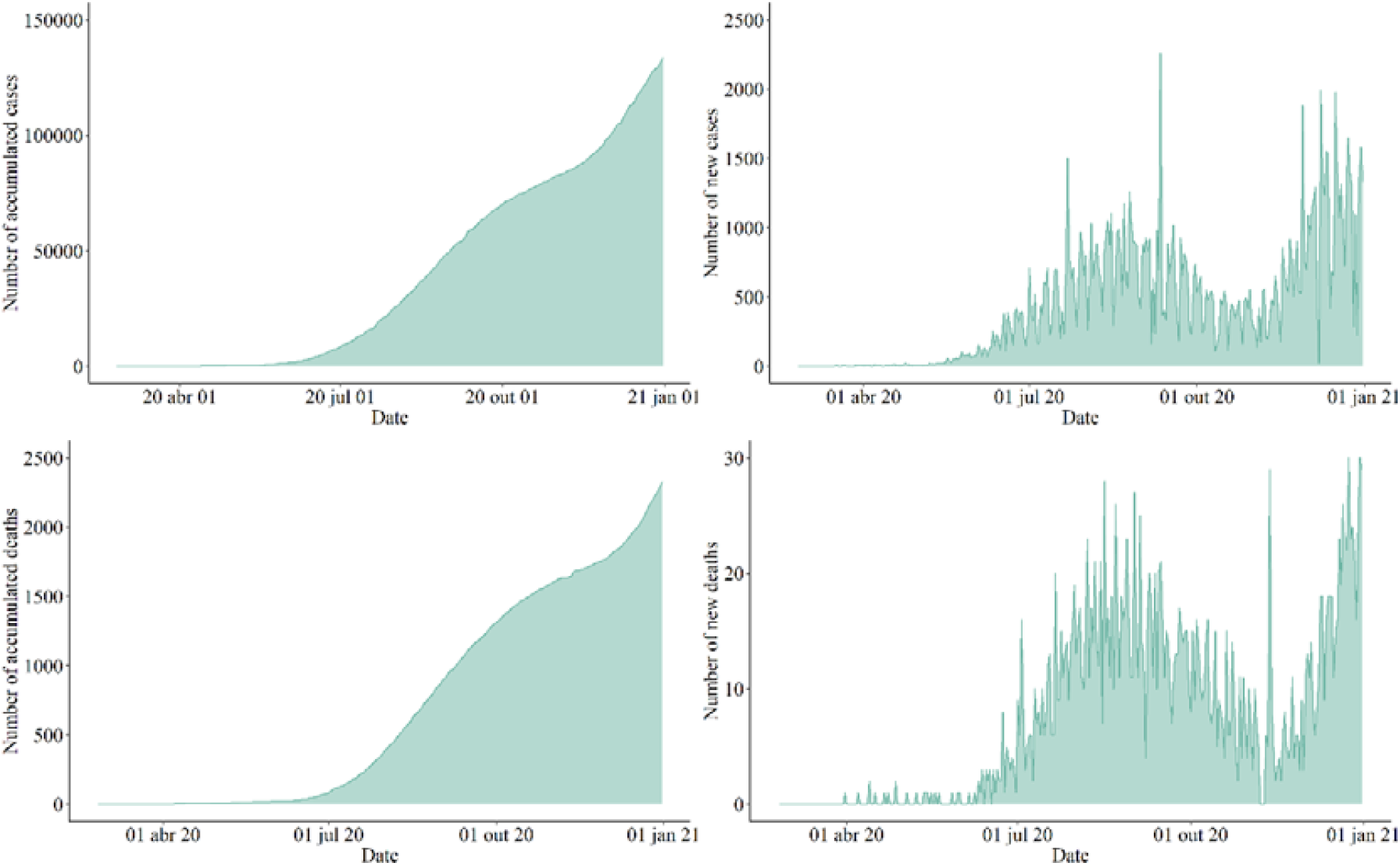
Number of new cases and cumulative cases of COVID-19 in Mato Grosso do Sul in the period from March 14, 2020 to December 31, 2020.

The moving average of the last 7 days, a measure adopted to verify the number of increase in COVID-19 cases and deaths in Brazil, showed a significant upward trend, both for new cases and for new deaths (Figure 2). The breaking point in the 7-day moving average series for new cases occurred on July 9, 2019, just before the breaking point in the 7-day moving average series for new deaths (Table 1).

**Table 1.**
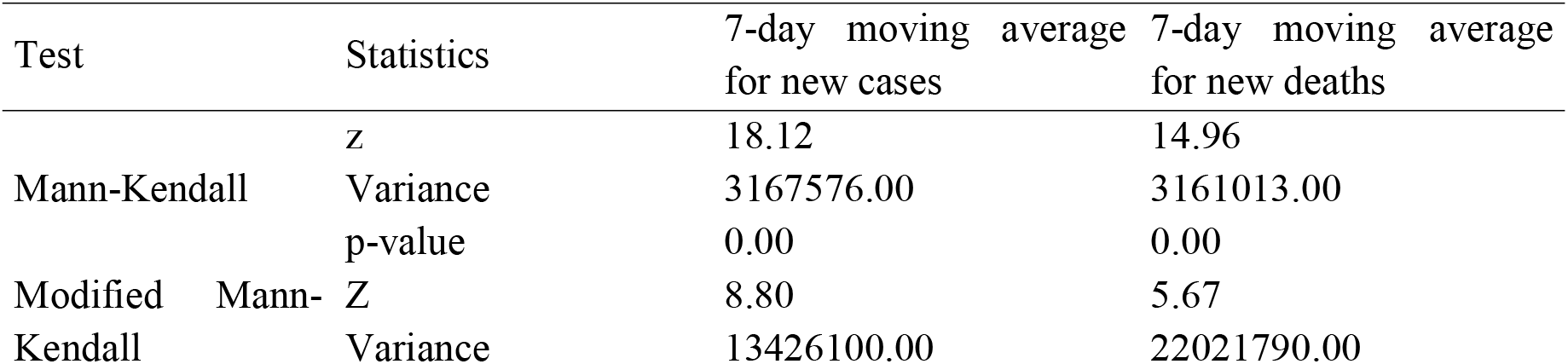
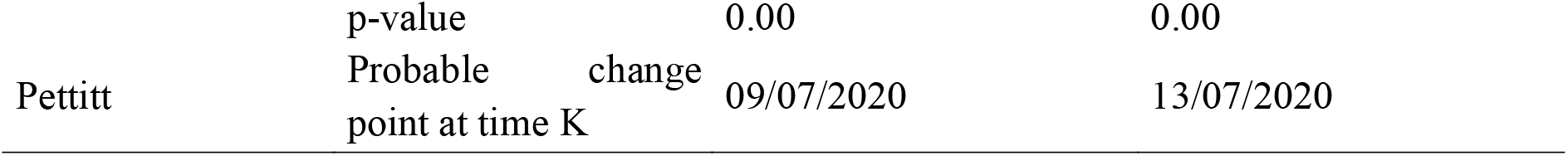
Non-parametric tests for verifying trends in the moving average of cases and deaths by COVID-19 in Mato Grosso do Sul from March 14, 2020 to December 31, 2020.

**Figure 2.**
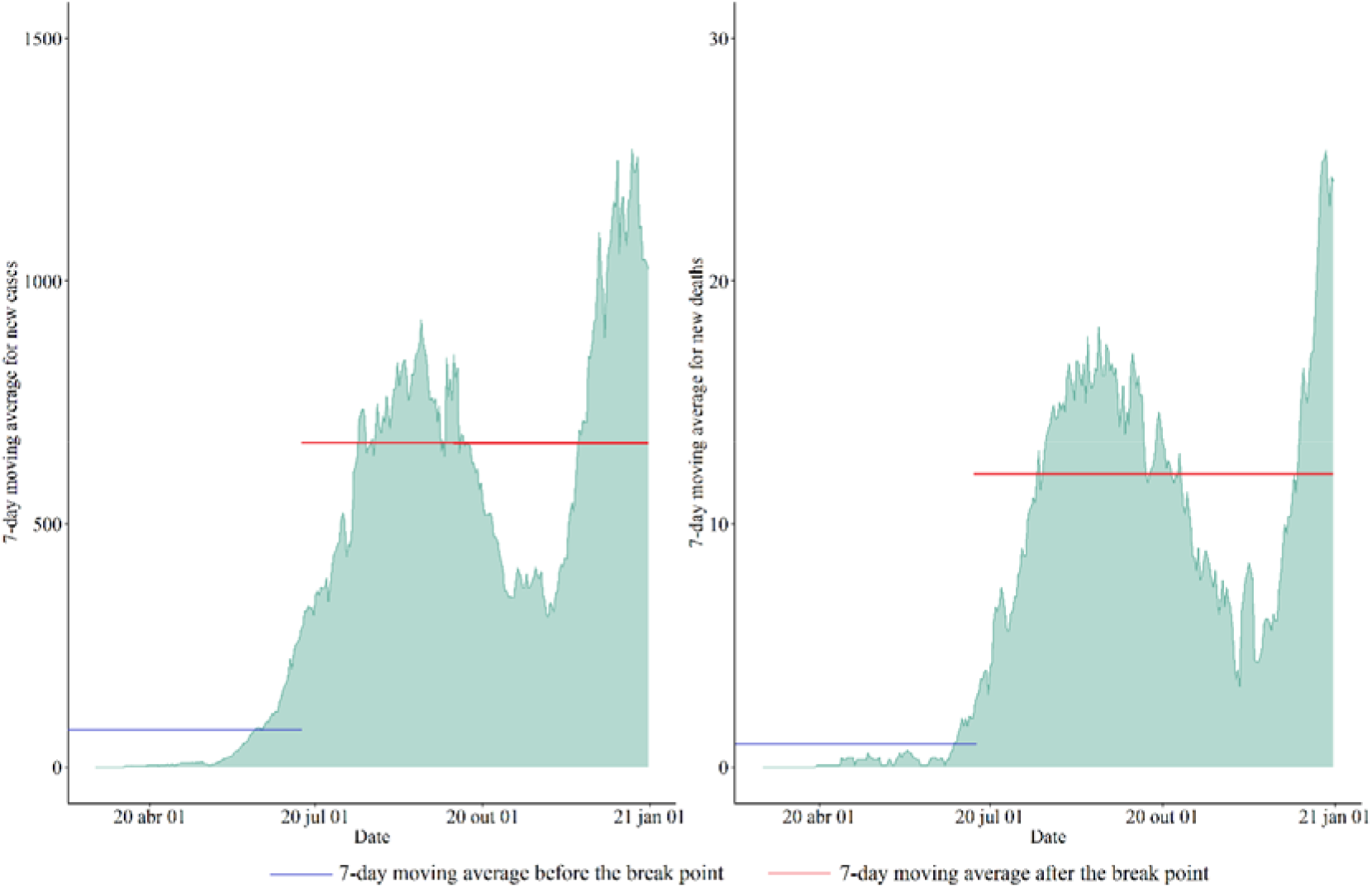
Moving average of the last 7 days for the number of new and new deaths due to COVID-19 in Mato Grosso do Sul in the period from March 14, 2020 to December 31, 2020.

There is a reduction in the 7-day moving average for MS, which showed a constant increase until reaching the first peak, at the end of September, decreasing in October, with new growth in November, until reaching the second peak, greater in magnitude than the first, in December. The trend tests for the moving average, used to monitor the number of cases, showed a significant upward trend, with a probable breaking point on July 9 and 13 for the number of new cases and the number of new deaths. There is a strong relationship (r> 0.90) between the 7-day moving average for the number of cases and number of deaths

The linear and non-linear regressions of the cumulative number of cases and deaths as a function of days were significant and the best fit was for the second degree linear regression and, in the case of the cumulative number of deaths, the exponential regression with 2 parameters (Table 2). The curvilinear shape of the total cases and accumulated deaths (Figure 3). The parameters β1 and β2 of the second degree linear regression represent the linear and quadratic effects, respectively, which shows that the accumulation of cases and deaths showed a higher rate of increase in the periods between July and October, with a slight decrease before a more expressive new rate, starting in November.

**Table 2.** Adjustment parameters and statistics for linear and non-linear regressions in Mato Grosso do Sul in the period from March 14, 2020 to December 31, 2020.

| Variables | Regressions | $\beta_0$ | $\beta_1$ | $\beta_2$ | $r^2$ | d | RMSE<br>(cases) |
| --- | --- | --- | --- | --- | --- | --- | --- |
| Accumulated<br>number of<br>cases | RL1 | -28535.68 | 435.61 | - | 0.90 | 0.85 | 12796.18 |
|  | RL2 | -1969.00 | -73.65 | 1.63 | 0.99 | 0.94 | 5028.47 |
|  | RE1 | - | 0.00 | - | 0.57 | 0.67 | 35833.57 |
|  | RE2 | 5018.00 | 0.01 | - | 0.95 | 0.87 | 9701.79 |
| Accumulated<br>number of<br>deaths | RL1 | -528.54 | 7.90 | - | 0.89 | 0.84 | 246.69 |
|  | RL2 | -48.04 | -1.31 | 0.03 | 0.97 | 0.92 | 124.74 |
|  | RE1 | - | 0.03 | - | 0.68 | 0.74 | 523.98 |
|  | RE2 | 0.01 | 92.98 | - | 0.93 | 0.84 | 211.81 |

RL1 = first degree linear regression; RL2 = second degree linear regression; RE1 = exponential regression with 1 parameter; RE2 = exponential regression with 2 parameters.

**Figure 3.**
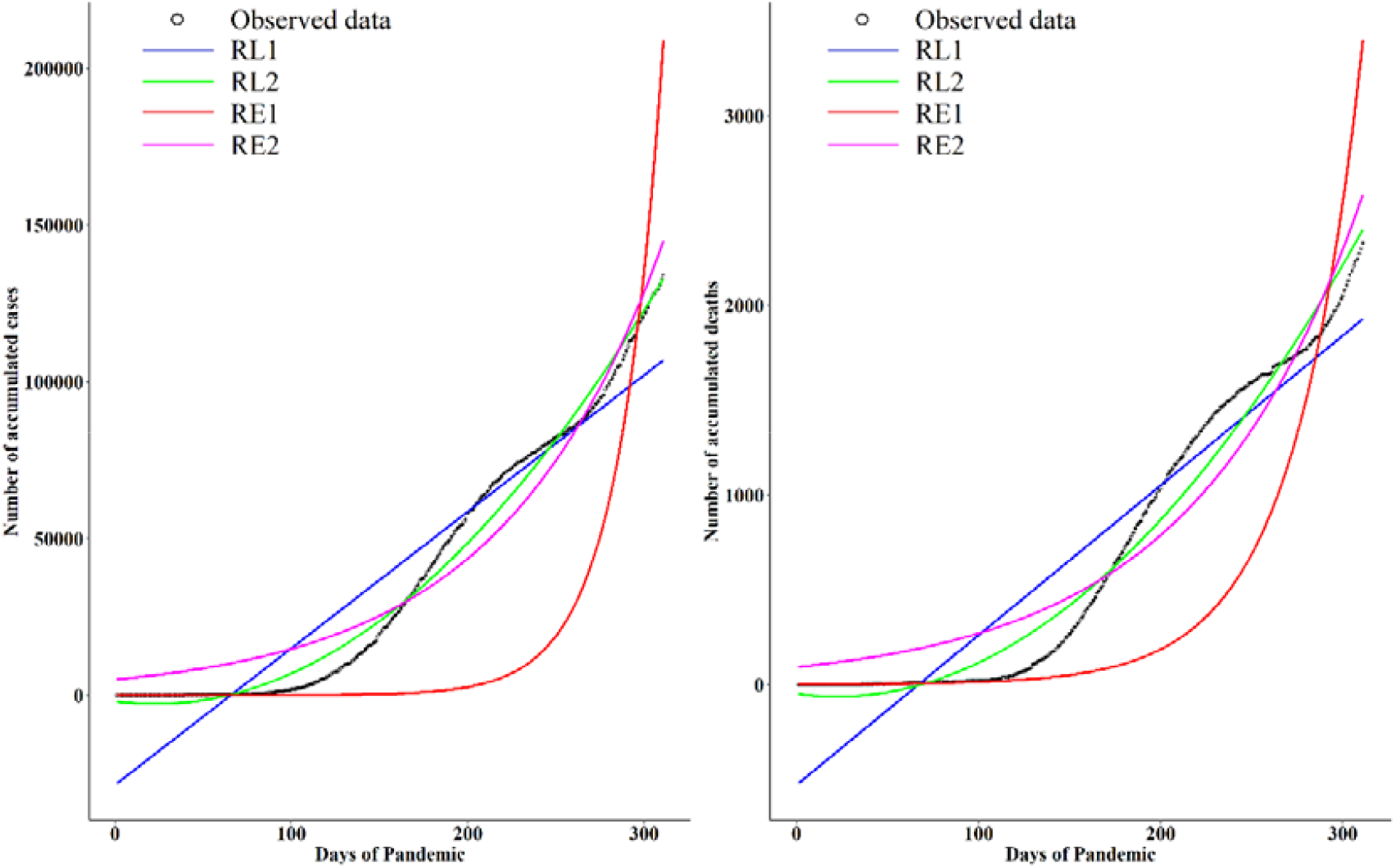
Linear and non-linear regressions for temporal characterization of the number of accumulated cases and number of accumulated deaths in Mato Grosso do Sul in the period from March 14, 2020 to December 31, 2020.

From the second week of May 2020, there was an important increase in the number of cases of covid, mainly for cases confirmed by the disease, with periods of growth until 07/22 where it reaches a value of 1503 cases, after which there is a decrease in 29 / 07 to 968 cases, growing again until 08/22 with 1177 cases, decreasing art 11/11 to 138 cases, growing again until 12/31 with 1220 cases. For deaths, there was also variation, but less. The highest growth rates occurred mainly in the first half of the analyzed period (08/16) with 28 deaths, then there was a fall on 11/13 to 11/30, with greater stabilization in this period, growing again on 12/2 and reaching the highest value of deaths on 12/24 and 12/30 with 30 deaths.

On average, the incubation period is estimated at five to six days, and can vary from zero to 14 days (BRASIL, 2020) (37). Clinical criteria and case definition can be performed in a clinical and laboratory manner. The laboratory diagnosis for the identification of the SARS-CoV-2 virus is carried out using real-time RT-PCR techniques (considered the gold standard for the identification of the new coronavirus) or rapid serological test validated by reference institutions (BRASIL, 2020) (37).

The spatial distribution of the incidence coefficient of COVID-19 in the municipalities of the state of MS demonstrated inequality in the incidence coefficient of this pandemic with spatial dependence and positive correlation associated with the MHDI, with the formation of a cluster in the municipalities close to the metropolitan region of Campo Grande (RMCG). Thus, the present study confirmed the association between COVID-19 and human development, pointing out the importance of geographic tracking in areas of potential local infectious transmission as a fundamental aspect to better coordinate actions to combat the pandemic.

Four clusters were identified for the state of MS (Figure 4), with the cities of Campo Grande and Japorã constituting clusters with a single element. The city of Campo Grande is the capital of MS, with the largest population (906,092 inhabitants), while the city of Japorã has 9,243 inhabitants. Campo Grande had the highest contamination rate by COVID-19 and a higher number of deaths, which are the reasons for forming a single group. The city of Japorã presented the highest lethality and the lowest COVID-19 contamination rate in the state, which are the reasons for forming individual clusters.

**Figure 4.**
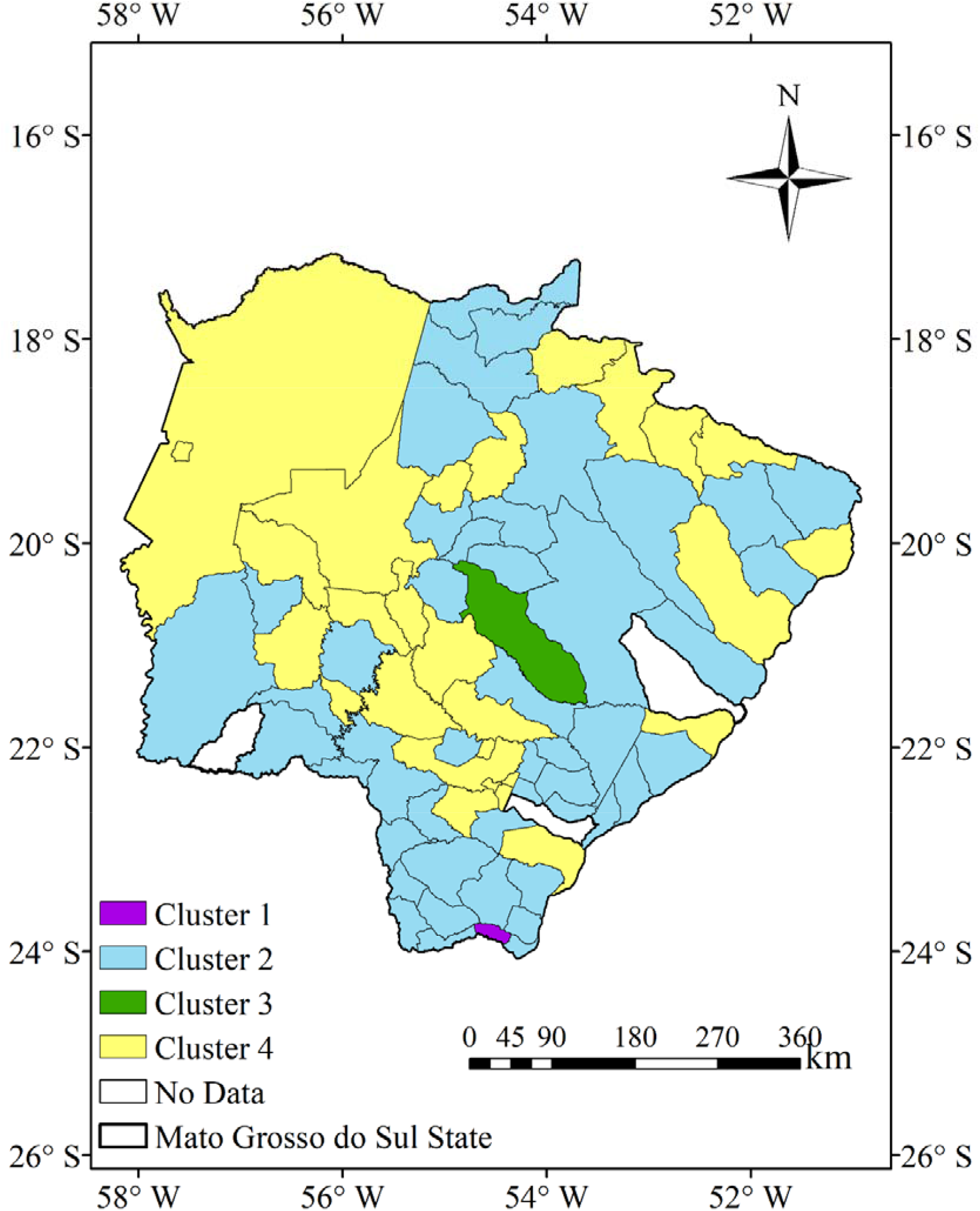
Clusters for the state of Mato Grosso do Sul, for the variables number of deaths, mortality, COVID infection rate, lethality and average HDI between March 14, 2020 to December 31, 2020.

Groups 2 and 4 showed a relatively well-defined spatial pattern. Group 2 (G2) - formed by a greater number of municipalities, which are located in the north-northeast, south-southeast, part of the central and southwest regions of the state. Group 4 (G4) consists of municipalities in the Pantanal region of southern Mato Grosso and central region of the state, in addition to some municipalities in the north and northeast, bordering the states of Mato Grosso (MT) and São Paulo (SP). Lethality was highlighted in group 1 (G1) due to its higher value in the state, formed by the city of Japorã, while for the variables mortality, deaths, contamination rate by COVID-19 and average IDH, the lowest value was observed for the state of MS. The G2 group presented intermediate values for all variables, being closer to the G1 group, with lower values of death, mortality and average IDH. The city of Campo Grande, which is part of the G3 group, had the highest average IDH and the highest contamination rate by COVID-19, number of deaths and mortality, while lethality was the lowest in the state of MS. The G4 group presented intermediate averages for all variables, being closer to the values observed in Campo Grande (group G3), with the exception of lethality. The CA technique was efficient in determining similar clusters in the state of MS, which can benefit the state in terms of public policies and actions to reduce the circulation of the virus and to mitigate the effects of the pandemic in the state. Since the technique was applied in other populous countries and with continental extensions (Holh et al., 2020))17) and even continents (Shariati et al., 2020) (29) with satisfactory results.

Figure 5 shows the boxplot analysis of slits of the formed clusters in which it is possible to notice the differences between them. It is possible to notice that at least one of the clusters showed significant differences in the median for the average HDI, COVID-19 infection rate, number of deaths, lethality or mortality. Baqui et al. (2020) (38) found several other important differences and sociodemographic trends specific to Brazil.

**Figure 5.**
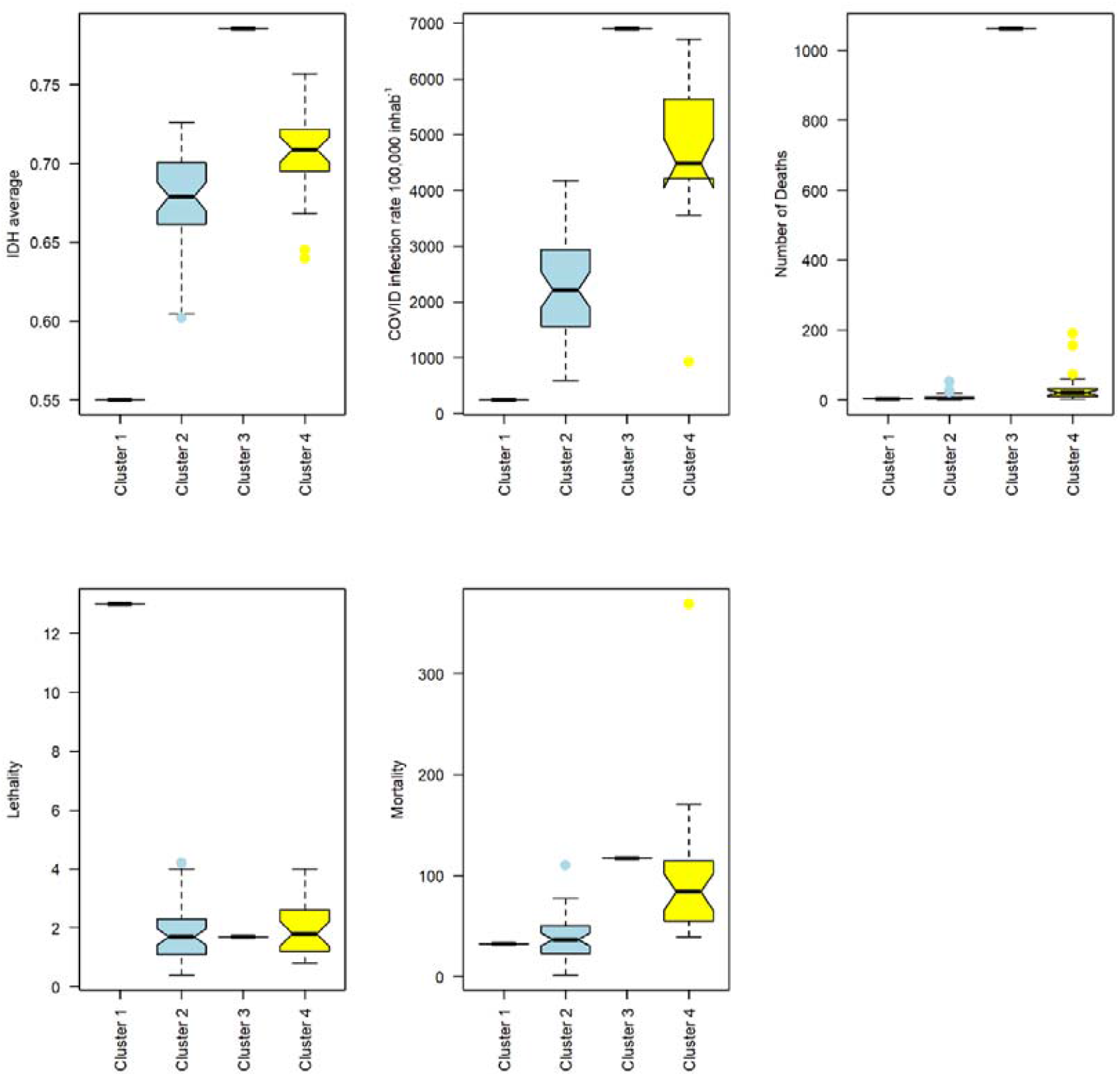
Crevice boxplot analysis for the mean HDI variables, COVID infection rate, number of deaths, lethality and mortality between March 14, 2020 to December 31, 2020.

The correlation between the following variables: number of deaths, mortality, COVID infection rate, lethality and average IDH between March 14, 2020 to December 31, 2020. Figure / Table 6/3 shows the analysis of the correlation between the Average IDH, COVID-19 infection rate, number of deaths, lethality and mortality. In general, the correlations were low with emphasis on the positive correlations between average IDH and COVID-19 infection rate, deaths and COVID-19 infection rate and deaths and average IDH. The significant negative correlation was between lethality and the average IDH.

|  | IDH<sub>average</sub> | TIC/100mil hab<sup>-1</sup> | Deaths | lethality | Mortality |
| --- | --- | --- | --- | --- | --- |
| IDH<sub>average</sub> | 1 |  |  |  |  |
| TIC/100mil hab<sup>-1</sup> | 0.46 | 1 |  |  |  |
| Deaths | 0.36 | 0.37 | 1 |  |  |
| lethality | -0.31 | -0.26 | -0.01 | 1 |  |
| Mortality | 0.30 | 0.38 | 0.21 | 0.25 | 1 |

The Barlett test (χ2 = 96.18 and p-value <0.05) and the KMO index (KMO = 0.55) indicated that the PCA could be used as a statistical tool to explain the variance of data related to the pandemic of COVID-19 in the state of MS. The first two main components (PC1 and PC2) explain 67.4% of the data variance (Figure 7). PC1 explains 42% of the data variance and represents a comparison between lethality and other variables (transmission rate by COVID-19, mortality, deaths and average IDH). The main variables in this component are: the transmission rate by COVID-19 and the average IDH. PC2, with 25.3% of the explained variance, shows the relationship between the average IDH and the variables lethality, mortality and deaths. In general, lethality and mortality have a greater influence on CP2.

**Figure 6.**
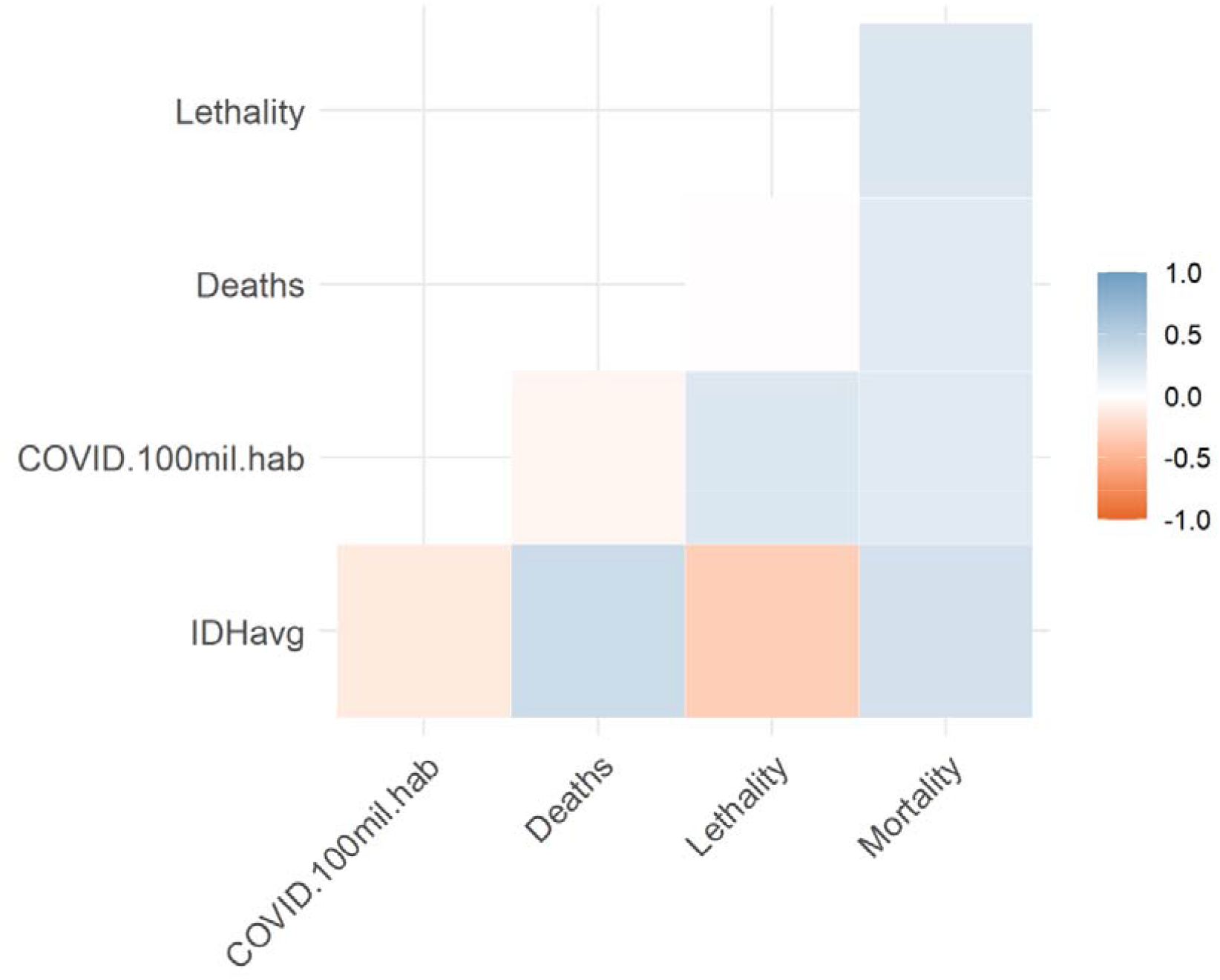
Correlation between the following variables: number of deaths, mortality, COVID-19 infection rate, lethality and average HDI between March 14, 2020 and December 31, 2020.

**Figure 7.**
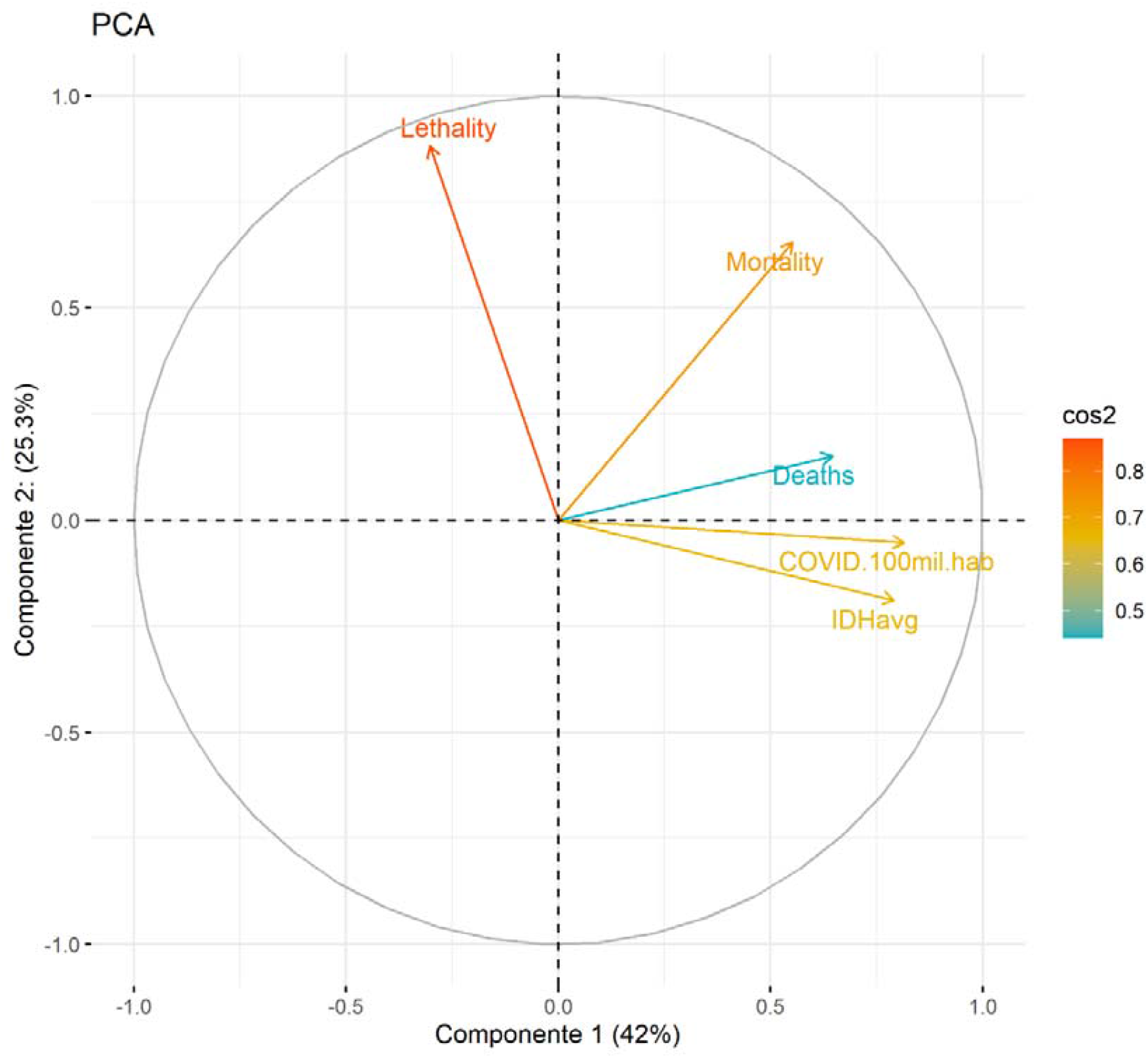
Graphical analysis of the variables number of deaths, mortality, COVID infection rate, lethality and average IDH in the first two main components.

The arrangement of the groups (Figure 8) obtained via the CA technique confirms that lethality is the main problem for group G1 (Japorã) and that group 2 presents intermediate results, with the second best situation in the state of MS for the following variables: rate transmission by COVID-19, mortality, deaths, lethality and average IDH. The G3 group (Campo Grande) had the worst results in terms of the pandemic and the G4 group had intermediate results, with the second worst situation in terms of the pandemic.

**Figure 8.**
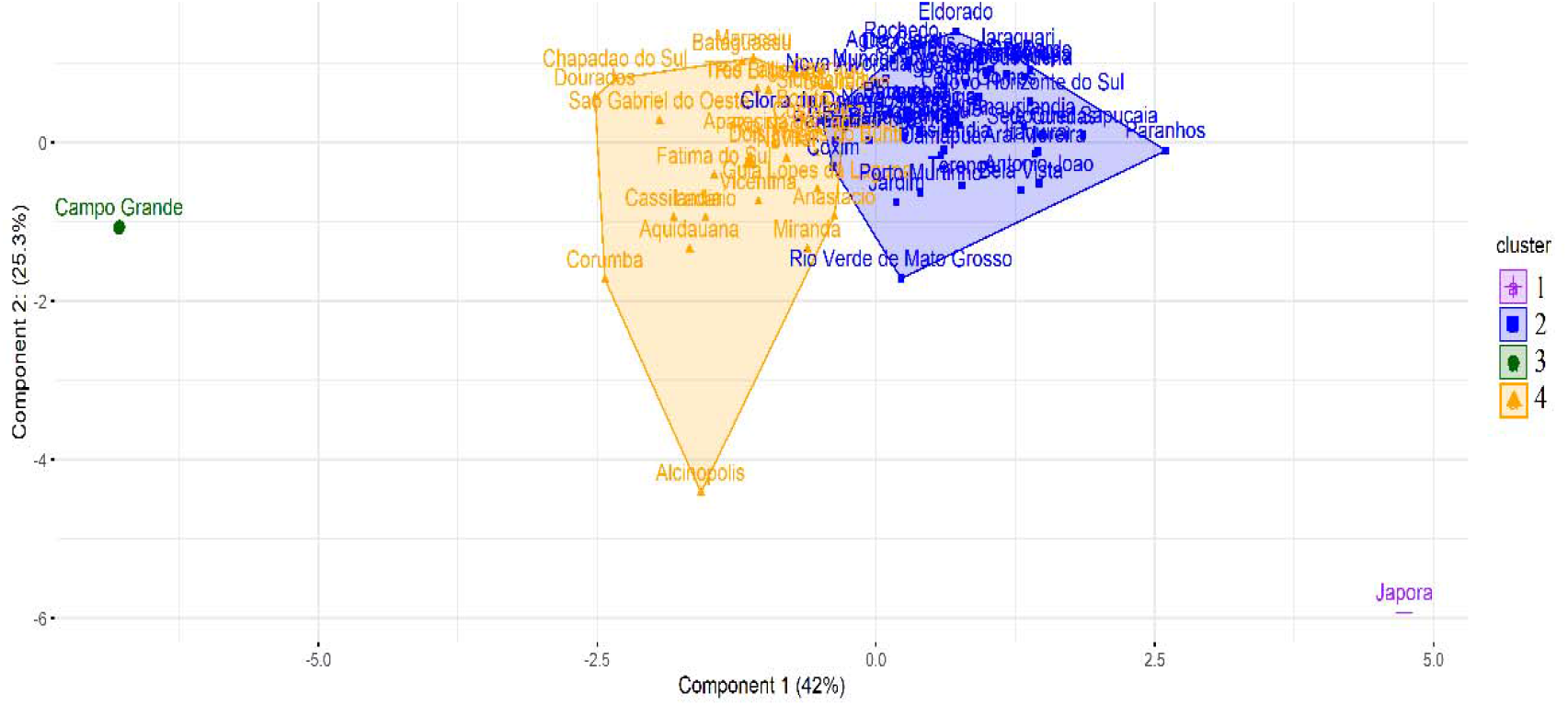
Components of the groups formed in the state of Mato Grosso do Sul and the first two main components.

## Discussions

The state of MS adopted social isolation measures at the beginning of the pandemic, at the end of March, which reflected in an isolation rate of over 50% by the end of April. In May, the isolation rate was less than 50%, which reflected an increase in the number of cases and deaths resulting from COVID-19. The rate of increase in the number of cases declined slightly at the end of October, although the isolation rate remained around 37% (INLOCO, 2020) (39). It is evident that only at the beginning of the pandemic in the state of MS, the contingency of the virus was effective. Reports in which the increase in COVID-19 cases in Brazil occurred since April are extensive (Marson and Ortega, 2020) (40).

In the case of Covid-19, there is a need for a critical assessment of epidemiological data related to human mobility to understand the dynamics of virus transmission at the local, regional and global scales. It is worth mentioning that epidemiological data differ between countries (Marson and Ortega, 2020) (39) and in Brazil between states when associated with sociodemographic data (Baqui et al. (2020) (38). The continuous integration of these data flows helps to guide the implementation resources to mitigate the transmission of COVID-19 (Candido et al, 2020) (41). However, the social isolation index and analyzes of the number of new cases and new deaths are strong evidence of the lack of commitment to mitigation of the effects of the pandemic Restrictions of social isolation is a recognized preventive measure for controlling the circulation of COVID-19 and studies have shown that the spread of the virus can be significantly reduced by this measure (Atalan, 2020) (42).

The positive spatial correlations found between the Covid-19 incidence coefficient and the measured social development, although weak, are unprecedented in Brazil, which can also be limiting from the perspective of comparing a few initial works on the topic addressed. In general, epidemiological research on communicable diseases finds in the average HDI an indirect indicator for health, since its greatest constitutive values are related to longevity, income and education (Andrade et al, 2012) (43).

The upward trends in the moving average of COVID-19 cases and deaths by COVID-19 are worrying and tend to reach worrying peaks due to the holiday season, coupled with holidays and school breaks, in which the population tends to travel and expand social relationships. The values of new cases and deaths represented by the 7-day moving average showed values not previously observed, which worries about the consequences on the health system of the Ministry of Health in the beginning of the year 2021, since the consequences of these peaks occur days after the diagnosis of the case. As Brazil does not have a coordinated national public health policy, the expected situation is catastrophic (Baqui et al., 2020; Fortaleza et al., 2020; Morato et al., 2020) (38; 44;45).

The understanding of the spatial distribution of COVID-19 in the MS, given its high magnitude of morbidity and mortality, the absence of effective immunization available and the pandemic character, which has not yet reached its maximum growth peak. Georeferencing proves to be a useful tool for epidemiological surveillance of communicable diseases and association with social determinants, in micro or spatial macro-analysis (Ribeiro et al., 2020) (46).

From a socioeconomic perspective, it is important to note that the elderly population represents one of the groups most subject to infection and symptomatology by COVID-19, with a higher risk of this cause of death for men. The pandemic caused by Covid-19 put the metropolises on alert, especially the large ones, which have high densities and facilitate the spread of the disease, as can be seen in the high incidence coefficient in Campo Grande and surrounding municipalities and in the municipalities that receive tourists as Bonito and Corumbá, with a tendency to diffuse less concentrated in the other municipalities of the state, although the measures taken by the state management contribute to limit the speed of occurrences, since the MS had an explosion of cases of Covid-19 in July (Figure 2).

The first cases of contamination occurred in the city of Campo Grande with the best MSI in MS and all those infected had made a trip abroad. The virus is already circulating on the outskirts of the city, with areas of high population density and worse sanitary conditions, which increases the concern with the speed of contagion. The metropolitan region of Campo Grande generates income and services, a tourist hub, has a high population density and urban mobility. This set of factors may corroborate the present study, suggesting that a greater IDHM may also facilitate the conditions of intense viral circulation, transmissibility and the increase in the clinical picture of COVID-19.

The low levels of IDHM reveal not only the population’s vulnerability, but also difficulties in health services regarding the diagnosis and treatment of the disease in the municipalities of Mato Grosso do Sul, similarly to the expected fragility of health services in Brazil (Ribas et al, 2019) (47) and in Latin American countries in tackling the pandemic (Rodrigues-Morales et al, 2020) (48). Given the great mobility of the population and the intense social and economic relations that the municipalities of the interior have with their respective capitals, the dissemination of Covid-19 should strongly affect the infrastructure in the interior. Unlike other illnesses at other times, this time there may not be enough time to transfer to the capitals or the capacity to meet all the demands of critically ill patients, which may reflect on different coefficients of lethality associated with social inequalities. In the same way, there is concern with the maintenance of jobs, since the weakening of employment ties was already underway because of the context of the current economic crisis in Brazil, but it is being exacerbated by the pandemic. In this sense, the number of people invisible to social policies, especially the homeless population, tends to increase, and ways of protecting the whole society against the new coronavirus need to be rethought. Despite the evolution of the Brazilian public health system, it is noteworthy that neglected diseases or those associated with health care in low-income populations can emerge secondary to COVID-19 and have an even more bleak impact on their cure prognosis. The study is valid in demonstrating the need for the articulation of epidemiological surveillance services in private health services with the SUS, the first being the most sought after by population groups with greater purchasing power. The arrangement of the groups obtained in the cluster analysis confirms that lethality is the main problem in group 1 (G1) (Japorã) and that group 2 (G2) presents intermediate results, with the second best situation in the state of Mato Grosso do sul for the variables: COVID transmission rate, mortality, deaths, lethality and average IDH. Group 3 (G3) (Campo Grande) had the worst results in terms of pandemic and group 4 (G4) intermediate results, with the second worst situation in terms of pandemic. Recent literature has focused on the generation of evidence in view of the following factors influencing the outbreak of COVID-19: endemicity for other infections, immune regulation with other co-infections, vaccination of the Calmette-Guérin bacillus (BCG), age of the population and quantitative data transposable for epidemiological predictions (Freitas e Silva, Pitzurra, 2020) (49).

## Conclusion

It is concluded that the occurrence of the Covid-19 incidence coefficient is unevenly distributed in the municipalities of the state of MS and was associated with the IDHM. The CA technique, together with the uneven Covid-19 mapping and its relationship with the IDHM in the Ministry of Health, can contribute to actions to tackle the regional pandemic, especially in the second wave of the national pandemic. The results obtained in this exploratory study contribute to the knowledge about the epidemic process of COVID-19 in the state of MS - Brazil and, thus, opens new perspectives for future analyzes on the behavior and variability of the disease. It is noticed the significant influence of the degree of commercial opening, as well as the occurrence of municipal elections in all of Brazil, which should not have been held, followed by fraternizations and agglomerations for purchases in commerce, common in Brazil, in the indicators of the disease, mainly in the increase of cases and deaths. The influence of the low availability of tests, significant underreporting and lack of adequate and organized information about the control of the pandemic cannot be ruled out.

## Data Availability

1. I have the right to post this manuscript and confirm that all authors have assented to posting of the manuscript and inclusion as authors.
Yes
2. I confirm all relevant ethical guidelines have been followed, and any necessary IRB and/or ethics committee approvals have been obtained.
Yes
The details of the IRB/oversight body that provided approval or exemption for the research described are given below

